# Differentiation of Fungal, Viral, and Bacterial Sepsis using Multimodal Deep Learning

**DOI:** 10.1101/2023.04.10.23288378

**Authors:** Aaron Boussina, Karthik Ramesh, Himanshu Arora, Pratik Ratadiya, Shamim Nemati

## Abstract

Sepsis is a major cause of morbidity and mortality worldwide, and is caused by bacterial infection in a majority of cases. However, fungal sepsis often carries a higher mortality rate both due to its prevalence in immunocompromised patients as well as delayed recognition. Using chest x-rays, associated radiology reports, and structured patient data from the MIMIC-IV clinical dataset, the authors present a machine learning methodology to differentiate between bacterial, fungal, and viral sepsis. Model performance shows AUCs of 0.81, 0.83, 0.79 for detecting bacterial, fungal, and viral sepsis respectively, with best performance achieved using embeddings from image reports and structured clinical data. By improving early detection of an often missed causative septic agent, predictive models could facilitate earlier treatment of non-bacterial sepsis with resultant associated mortality reduction.

## Introduction

Sepsis is a major cause of morbidity and mortality around the world. In the United States alone, sepsis potentially accounts for nearly a quarter of a million excess deaths yearly^1^. Moreover, there are serious costs and other health burdens associated with each sepsis related admission for both patients and health systems^2^.

While the majority of sepsis is caused by bacterial pathogens, there is a significant volume of sepsis caused by fungal and viral agents. Some estimates suggest ∼20% of sepsis is caused by fungi. Furthermore, these less common classes of infective agents can often contribute to greater morbidity and mortality, with estimates for fungal sepsis mortality ranging from 40-60%^3^. However, current standard culture methodologies can result in serious delays to administration of appropriate treatment. The increased mortality from fungal sepsis thus can be potentially attributed to both a more acutely ill patient population as well as delayed identification of the causative agent^4^. All together, further efforts to more accurately and rapidly differentiate the causative microorganism class of sepsis could prove crucial to decreasing excess associated morbidity and mortality.

Previous work has established a number of patient risk factors contributing to greater likelihood of primary fungal sepsis most commonly including immunosuppression of various primary etiologies^5^. Additionally, there have been prior efforts to classify patients into viral, bacterial, and fungal etiologies of pneumonia based on machine learning interpretations of standard chest x-rays^6^. Finally, there has also been work differentiating laboratory test results between classes of sepsis^7^. While these efforts have all been conducted separately, there seems to have not yet been an effort to combine these disparate patient factors together to predict sepsis type.

As such, in this study, the authors aim to use a coordinated multi-task approach drawing from chest-xray images, radiology reports, and clinical lab data to differentiate between bacterial, fungal, and viral sepsis in the MIMIC-IV dataset.

## Methods

Predictive methods for differentiation of septic class were based on chest x-ray image, chest x-ray radiology report, and structured clinical data from the MIMIC-IV database^8^.

Labels for the infection class were taken from the microbiology events data provided in the main MIMIC-IV database. Culture results were grouped by patient and time point to develop time specific labels for bacterial, fungal, and viral infection based on isolated organism name. As such, at any given time point it is possible for a patient to have more than one positive infective class. Patients were labeled culture positive for one day before and seven days after the order time for a positive culture result.

Structured data on medications, vital signs, laboratory results, demographics, and other clinical elements was extracted directly from the MIMIC-IV database. Any patient with a microbiology result was included. Medications were processed from the prescription orders and mapped to 16 distinct classes including antibiotics, antivirals, antifungals, anticoagulants, and vasodilators. Six distinct vital signs (temperature, heart rate, respiratory rate, O2sat, systolic blood pressure, and systolic blood pressure) and one procedure (arterial line insertion) were extracted from the flowsheets. The full list of clinical variables is presented in Supplemental Table 1. All units from a patient’s encounter were included and dynamic variables were sampled and updated at hourly intervals throughout the encounter. Multiple measurements within a time bin were averaged and missing values were imputed using the last observation carried forward for 24 hours. The changes in measurement values between successive vital-signs and laboratory results as well as the number of hours since the last measurement were preserved as additional features. Sepsis labels were assigned to patients admitted to the ICU using a combined sepsis definition including Sepsis-3 and Sepsis-CMS.

Patient chest x-ray images were collected from the MIMIC-CXR-JPEG^9^ dataset and mapped to x-ray reports and structured patient data. Features were extracted from image data using a pre-trained CheXNet model^10^. CheXNet, is a 121-layer convolutional neural network based on DenseNet-121 that inputs a chest X-ray image and outputs the probability of pneumonia along with a heatmap localizing the areas of the image most indicative of pneumonia. CheXNet is trained on the ChestX-ray14 dataset, which contains 112,120 frontal-view chest X-ray images individually labeled with up to 14 different thoracic diseases, including pneumonia. Images from the MIMIC-CXR-JPEG dataset were resized, and then fed into the pre-trained model to extract x-ray image embeddings.

In addition to chest x-ray images, embeddings were also extracted from radiology reports using Clinical BERT^11^, a unique version of BERT pre-trained on 2 million notes from the MIMIC III database. Clinical BERT contains 12 transformer heads, and has a total of 110 million parameters. We concatenated together the text report into a single sequence and passed it to the model. Every sequence was padded to a 128 token sequence length after appending <CLS> and <SEP> tokens. The final transformer head of the model returned a 768 dimension embedding corresponding to each token in the sequence.

Using the aforementioned features, a neural network was trained to classify bacterial, fungal, and viral sepsis. The network architecture is described in Figure 3 and consists of 3 hidden layers and 3 output nodes. The network outputs separate predictions for each category and allows for multiple labels. Two of the hidden layers are shared between the multiple classification tasks to enable the network to learn common representations. ReLU activation is used for all of the layers except for the output nodes which use sigmoid activation. The network was trained using Adam optimization with a learning rate of 0.005. The cost function was the average binary cross entropy loss across all three output nodes and training occurred for 100 epochs with early stopping. Model performance was evaluated on a 20% test set. Three separate experiments were run using these hyperparameters. First, the performance of the model was evaluated using only the 133 clinical features. Next, the performance was evaluated with the inclusion of the radiology report embeddings. Finally, the model was evaluated using all of the clinical features, report embeddings, and chest X-ray embeddings. The model was trained on the overall cohort and evaluated on septic patients admitted to the ICU.

**Figure 1:**
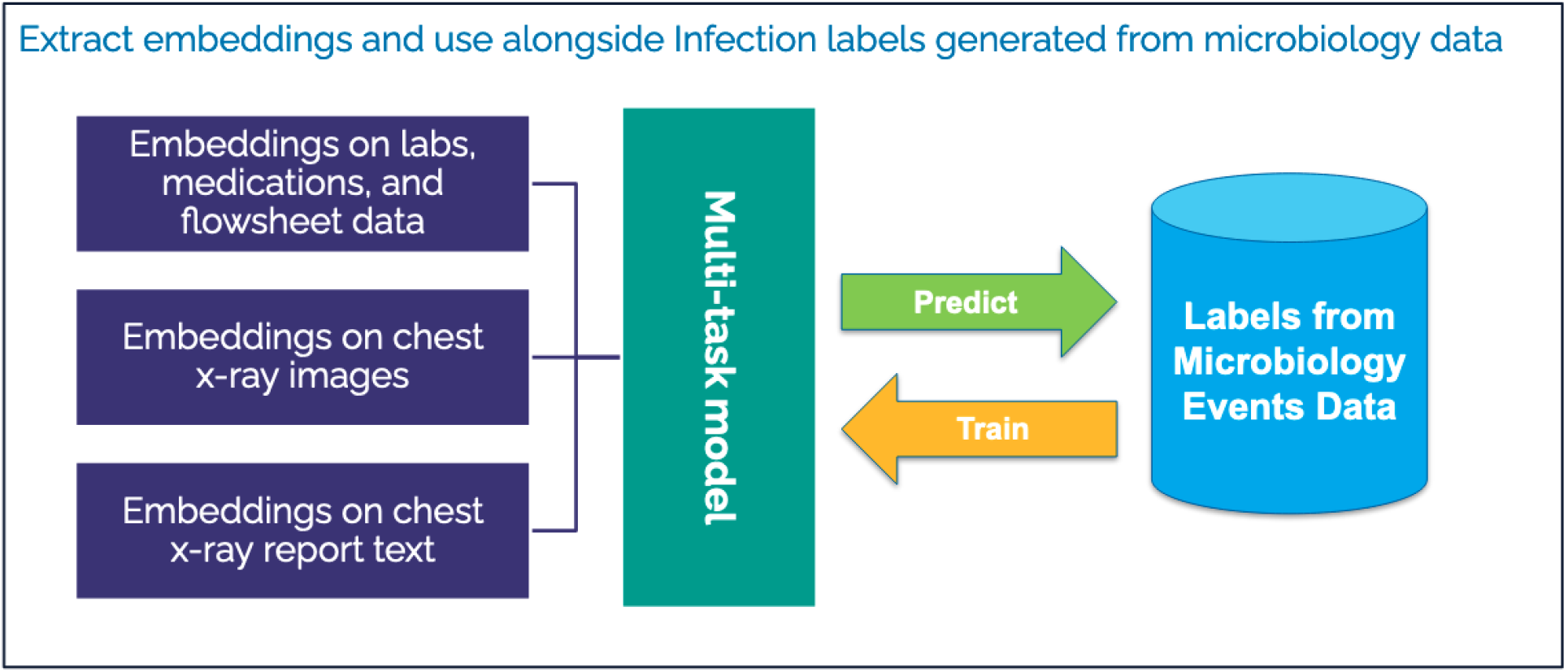
Graphical representation of feature extraction and embedding generation from MIMIC-IV inpatient database)

**Figure 3:**
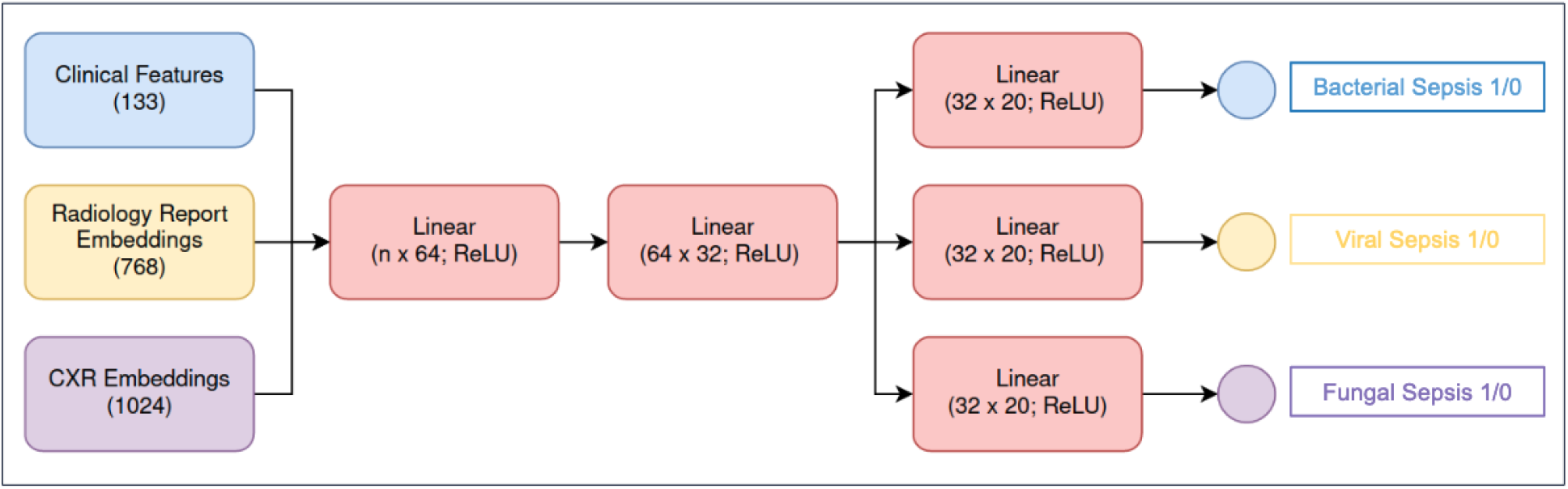
Graphical representation of neural network used to generate multi-task predictions.

**Figure 4:**
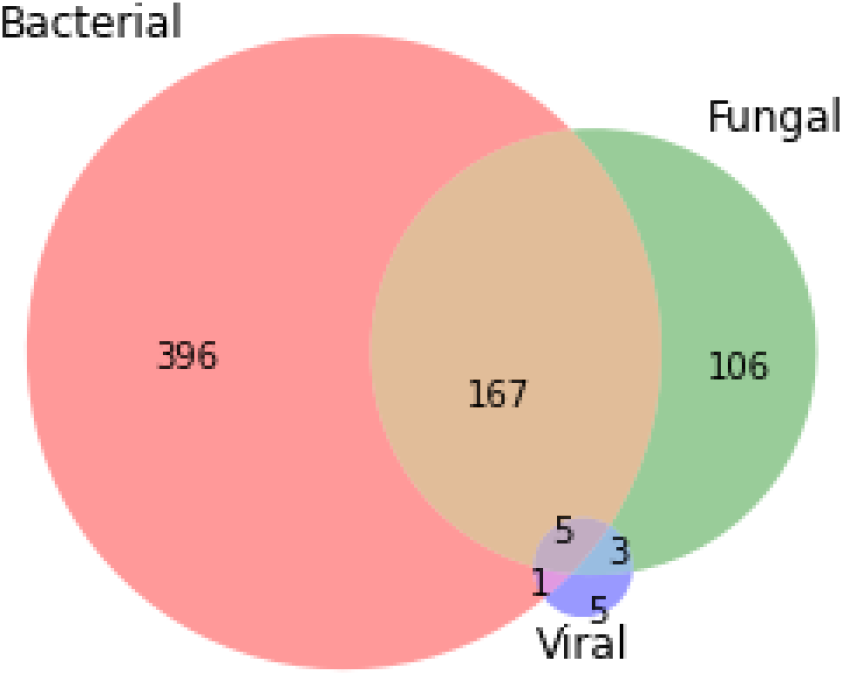
Venn Diagram representation of overlapping septic labels showing total number of septic patients per class.

## Results

The final cohort used for analysis is described in Table 1. The dataset consists of 170,624 encounters, 25.6% of which had positive bacterial cultures, 6.3% of which had positive fungal cultures, and 0.6% of which had viral infections.

**Table 1.** Patient characteristics across microbiology results.

| Metric | Overall Dataset | All Culture Positive | Bacterial Positive | Fungal Positive | Viral Positive |
| --- | --- | --- | --- | --- | --- |
| Patient Count, N (%) | 170,624 (100%) | 48,695 (28.5%) | 43,642 (25.6%) | 10,725 (6.3%) | 1,021 (0.6%) |
| Age, Median (IQR) | 62 (49 - 74) | 64 (51 - 76) | 63 (51 - 75) | 65 (53 - 76) | 61 (49 - 73) |
| Female, N (%) | 85,397 (50%) | 24,305 (50%) | 21,560 (49%) | 5,580 (52%) | 502 (49%) |
| Hospital Stay (Days), Median (IQR) | 4.46 (2.2 - 8.1) | 7.08 (3.7 - 13.5) | 6.95 (3.7 - 13.4) | 12.46 (6.5 - 22.3) | 7.5 (3.4 - 17.6) |
| Sepsis Patients, N (%)* | 1,138 (100%) | 683 (60.0%) | 569 (50.0%) | 281 (24.7%) | 14 (1.2%) |

\* Percentages are expressed relative to the septic subpopulation and patients may appear in more than one class

The model performance on the septic subpopulation test set for all three experiments is presented in Table 2. The highest performing model included the clinical features as well as the radiology report embeddings from ClinicalBERT.

**Table 2.** Results on the study septic population test set.

|  | **Bacterial AUC** | **Viral AUC** | **Fungal AUC** |
| --- | --- | --- | --- |
| **Clinical Features Only** | 0.71 | 0.76 | 0.82 |
| **Clinical Features +**<br>**Note Embeddings** | 0.81 | 0.79 | 0.83 |
| **Clinical Features +**<br>**Note Embeddings +**<br>**Image Embeddings** | 0.80 | 0.79 | 0.82 |

## Discussion

In this work, the authors developed a model that achieves fair classification performance on identifying the infectious organism class in septic patients. To the authors’ knowledge, this is the only work to-date that has incorporated the multimodal data elements available in MIMIC IV for the purpose of infectious organism classification. A central finding of our work is that the inclusion of unstructured clinical text from the radiology report improves the classification performance, particularly for cases of bacterial infection. Interestingly, there was no additional utility from the inclusion of CheXNET embeddings on the source chest X-ray images. This suggests that under current methods, radiology reports contain the necessary information for differentiation of septic class, highlighting the value of this data source for future efforts.

As expected, there was an increased length of stay for fungal culture positive patients. This aligns with established data suggesting fungal infections in septic patients represent largely nosocomial origins. Additionally, there is significant overlap between some classes of septicemia, suggesting that there are layered infectious hospital courses for patients who develop sepsis. However, in current efforts, there were no labels specified based on culture site, organism resistance, or if the culture was conducted after a lengthy inpatient stay. As such, it is difficult to quantify the severity of inpatient fungal infections accurately. Future work could potentially investigate infections of pulmonary origin alone, and include mortality data for improved label creation.

As established in previous literature, there seem to be clear clinical factors which precipitate fungal infection, such that image data or radiology reports are not necessary to identify patients at high risk. Neutrophil count, C reactive protein, procalcitonin, and other laboratory measures have been cited previously as being important markers of fungal septicemia. However, this model is potentially limited by significant missingness of such measures. Moreover, there is a risk that the presence of such measures alone helps to indicate a septic patient, leading to model bias. Future efforts may focus on training and validation of the model on only a population of septic patients, as opposed to a broader inpatient population.

Finally, while in this work labels were indicated positive for one day before and seven days after a positive culture result, the true clinical reality may not align with such labeling. Specifically, it is noted that signs of sepsis are often detectable in a highly variable time period ahead of culture positivity. Additionally, fungal septicemia often occurs in those patients already admitted to the hospital with further associated delays due to prolonged fungal culture timelines.

## Conclusion

Fungal and viral sepsis are underdiagnosed and often superimposed varieties of sepsis with serious associated morbidity and mortality. While some efforts have been made to highlight clinically detectable differences between sepsis classes to facilitate early detection of non-bacterial sepsis, none have thus far incorporated both structured clinical data and relevant imaging. Using such a combined model, fair performance is possible on the task of sepsis class identification, with improved performance most notable through additional information types in bacterial sepsis detection. Such efforts could speed treatment of non-bacterial sepsis, reducing sepsis mortality. Future work on the matter of non-bacterial sepsis detection may include greater sophistication in assigned sepsis labels, experimentation with culture positivity allowance, and further model architecture improvements.

## Data Availability

All data produced in the present study are available upon reasonable request to the authors

## Acknowledgements

S.N. is funded by the National Institutes of Health (R35GM143121). He is co-founder of a UCSD start-up, Healcisio Inc., which is focused on commercialization of advanced analytical decision support tools. Mr. Boussina is funded by the National Library of Medicine (#2T15LM011271-11). The opinions or assertions contained herein are the private ones of the author and are not to be construed as official or reflecting the views of the NIH or any other agency of the US Government.

## Supplementary Tables

**Supplementary Table 1.** Clinical variables extracted from the structured MIMIC IV data. Missingness is reported as the percentage of encounters without a measurement.

| Clinical Variable | Missingness (%) |
| --- | --- |
| Age | 0 |
| Gender | 0 |
| Length of Stay | 0 |
| Care Unit | 0 |
| Medications | 0.1 |
| Hematocrit | 3.8 |
| Platelet Count | 5.6 |
| Hemoglobin | 5.8 |
| MCH | 5.9 |
| MCHC | 5.9 |
| Red Blood Cells | 5.9 |
| RDW | 5.9 |
| MCV | 6.1 |
| White Blood Cells | 6.2 |
| Creatinine | 7.2 |
| Urea Nitrogen | 7.9 |
| Chloride | 8.5 |
| Bicarbonate | 9.3 |
| Magnesium | 16.1 |
| Potassium | 16.5 |
| Total Calcium | 17.5 |
| Phosphate | 18.2 |
| Sodium | 20.1 |
| Anion Gap | 22.1 |
| PT | 41.5 |
| PTT | 43.6 |
| Alanine Aminotransferase (ALT) | 56.8 |
| Aspartate Aminotransferase (AST) | 57.0 |
| Bilirubin, Total | 57.5 |
| Alkaline Phosphatase | 57.7 |
| Basophils | 60.1 |
| Eosinophils | 60.1 |
| Monocytes | 60.1 |
| Lymphocytes | 60.1 |
| Neutrophils | 60.4 |
| pH | 63.8 |
| Albumin | 68.2 |
| Lactate Dehydrogenase (LD) | 74.5 |
| Red Blood Cell Differential | 75.7 |
| White Blood Cell Differential | 75.8 |
| Lactate | 79.3 |
| Base Excess | 82.3 |
| pCO2 | 82.8 |
| pO2 | 84.6 |
| Glucose | 85.5 |
| Ferritin | 90.8 |
| Transferrin | 91.0 |
| Fibrinogen | 91.7 |
| Oxygen Saturation | 92.1 |
| Bilirubin, Direct | 96.4 |
| Arterial Line Insertions | 91.7 |
| Sedimentation Rate | 97.3 |
| C-Reactive Protein | 98.0 |
| Ammonia | 99.2 |
| D-Dimer | 99.7 |

